# Clinical applicability of the WaveFront Phase Imaging Sensor in the Keratoconus Disease

**DOI:** 10.1101/2023.11.20.23298756

**Authors:** Carolina Belda-Para, Gonzalo Velarde-Rodríguez, Miriam Velasco-Ocaña, Juan M. Trujillo-Sevilla, Iván Rodríguez-Méndez, Javier Rodríguez-Martin, Nicolas Alejandre-Alba, Silvia Rodríguez-García, José M. Rodríguez-Ramos

## Abstract

**Significance:** The analysis of the wavefront phase of keratoconus eyes has become increasingly important due to its impact on the assessment of visual quality loss, irrecoverable with conventional optical treatments, and the understanding of morphological changes in corneal structures.

**Aim:** The aim of this work is to quantitatively assess the wavefront phase of keratoconic eyes measured by the novel high-resolution ocular aberrometer t·eyede (based on WaveFront Phase Imaging Sensor).

**Approach:** The measurements with this device were taken on healthy and keratoconic eyes at the Hospital Universitario Fundación Jiménez Díaz (Madrid, Spain). The wavefront phase recovered is post-processed and displayed in different phase maps, using exclusively the high-pass filter map for this study. From this map, the Root Mean Square, the Peak-to-Valley, and the amplitude value of the predominant obtained from its Fourier Transform are the parameters to distinguish between healthy and keratoconic eyes. Furthermore, the banding pattern observed in most keratoconic eyes is quantitatively analyzed by means of its period and orientation calculated from the Fourier Transform.

**Results:** The Control group was composed of 43 healthy eyes, and the Pathological group presented 43 keratoconic eyes. Regarding the first three parameters used to compare both groups, there were significant statistical differences between them in all parameters, where the values obtained with the keratoconic group were higher than those of the healthy group. Although the values cannot be statistically comparable between stages, they tended to increase according to the severity of the disease. On the other hand, analyzing the banding pattern, the mean value of the period was approximately 50 µm, and most orientations were obliquus, tending to be horizontal rather than vertical.

**Conclusions:** The present study demonstrates that the objective analysis of the high frequencies detected by this wavefront sensor is a potential tool to reliably advise in the keratoconus diagnosis. Furthermore, combining this achievement with the quantitative assessment of the banding pattern would be used to objectively monitor the internal corneal status throughout the disease and its treatment optimization and surgery time.

## 1 Introduction

Keratoconus is a progressive and noninflammatory asymmetric corneal disorder characterized by central corneal thinning and conical corneal shape. Its prevalence is about 0.5% in the general population^1^ and leads to irregular astigmatism, increased higher-order aberrations, and progressive vision loss^2–4^.

In general, Keratoconus is usually graded according to the standardized Amsler-Krumeich classification, which includes the parameters of keratometry and the thinnest point of the cornea, as the findings observed in the biomicroscopy exploration (corneal scarring, Vogt striae, predominant corneal nerves, etc.). Recently, some authors have brought the Zernike coefficients into relevance to objectivate this classification, concretely using the vertical coma coefficient^5,6^ due to the large amount of this polynomial induced by the formation of the conic protrusion. There are different techniques to obtain these values: by topographers (Placido’s disk, Schemiplflug camera, etc.) or by wavefront sensors (Hartmann-Shack, WaveFront Phase Imaging, laser ray tracing, etc.). Then, it is necessary to clarify that topographers extract Zernike polynomials using the elevation map of the corneal surface instead of using a spherical surface as reference (such as the wavefront sensors do), where a customized algorithm is required to apply to calculate them for each surface that depends on the manufacturer device^4^, obtaining a lack of repeatability for a highly deformed cornea (such as the keratoconic cornea) has been reported in the literature^7–9^. Meanwhile, wavefront sensors acquire the phase directly by refraction of light passing through all refractive ocular surfaces. Hence, the results obtained by both methods must not have been numerically comparable without using a data treatment. Although both acquisition methods are disparate and the plane measurement reference is different too (topographers use the cornea as the measuring plane while ocular aberrometers use the pupil plane), their comparisons could be made concerning their trends.

However, the evaluation of the keratoconic eyes by ocular aberrometers (especially those that use the Hartmann-Shack sensor) is limited to the 8th or 10th order of Zernike polynomials for different reasons. One of these is that further orders have not been considered clinically necessary to assess pathologies due to the low values obtained; another is the limited resolution of this sensor, which depends on its array of microlenses and its correspondent spots. At last, in moderate-severe cases, the conical surface deflects considerably the light, displaying the spots far from their reference on the image detector (CCD, CMOS), translated as phase distortion^10–12^.

To overcome this, the WaveFront Phase Imaging (WFPI) sensor has recently been presented in the literature to measure the phase in transparent objects with more resolution than Hartmann-Shack, being previously validated in other technical fields such as silicon metrology^13,14^, optical glass quality assessment^15,16^, and tested in ophthalmology^17^ with healthy subjects and keratoconic patients by t·eyede prototype^18–20^. In the last work, the authors have demonstrated that this device is capable of reliably distinguishing between healthy and keratoconic eyes by means of the ocular aberrations (Astigmatism, Coma, and High Order Aberrations), besides exposing the relevance of the further orders than 10th by revealing a pattern of folds or bands in advanced keratoconus that have not been seen in phase results previously. Therefore, the analysis of the known parameters combined with the quantitative assessment of these patterns could be helpful in the diagnosis, monitoring, or treatment of keratoconus.

Previously, the map that reveals these folds had only been submitted qualitatively but can be described quantitatively using the map itself and assessing it by means of the Fourier Transform (FT). This mathematical method has been applied to numerically evaluate several biological tissues featured by bands (as collagen structures) observed in other techniques^21,22^. Hence, due to the versatility of this imaging treatment, it should be applied to achieve the differences between healthy and keratoconic eyes and characterize the patterns obtained in those.

The aim of this study is to quantitatively assess the signs observed on the phase maps in keratoconus eyes against the healthy ones as well as numerically characterize the features observed in those keratoconic maps applying the Fourier Transform.

## 2 Materials and methods

### 2.1 Set-up: t·eyede aberrometer

The device presented in this document is the t·eyede aberrometer, which methodology has been previously described in other works^17,19,20^. This aberrometer can obtain the ocular phase using the WaveFront Phase Imaging sensor (Wooptix S.L., La Laguna, Tenerife, Spain), whose main advantage over other phase sensors is its lateral resolution of 8.6 µm. That fact is due to the possibility of using the pixels of an imaging sensor (such as CCD or CMOS) as measurement points without needing any other optical element to sample phase (like an array of microlenses or a pyramidal prism). The sensor captures two intensity images shifted at the same distance to either side of the pupil plane to recover the wavefront phase from a transparent sample^14,19^, as shown in the original Fig 1(a) in Bonaque’s work^19^.

**Fig. 1.**
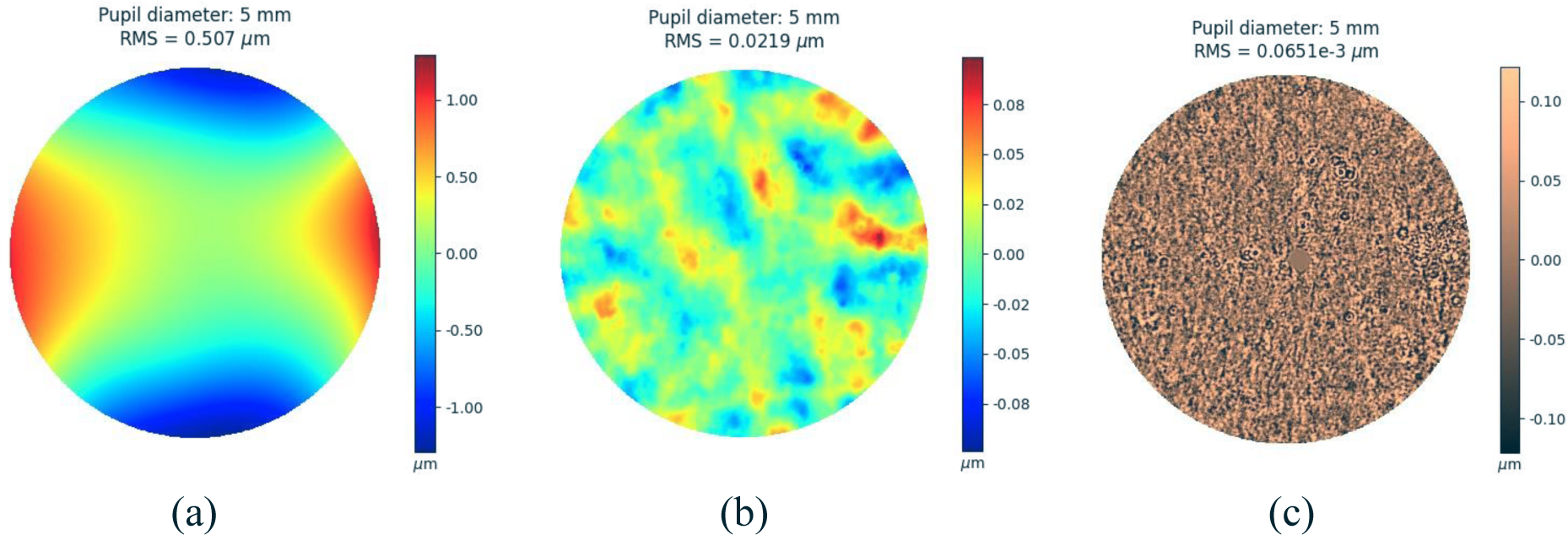
Healthy eye: (a) Low order phase map composed by the first 65 Zernike’s polynomials removing piston, tip/tilt, and defocus, (b) Extremely high order map obtained from the subtraction of the rough phase and the first 65 Zernike’s polynomials, and (c) High-pass filter map of the extremely high order map (b).

Regarding the apparatus, it uses a Superluminescent Diode (SLD) with a wavelength of 780 nm. The exposure time in the pupil plane is 30 ms at a power of 0.78 mW, which is under the security value of the ANSI rules. A Badal system corrects the patient’s defocus in the range of -10.00 D to +10.00 D. Finally, the CCD camera captures two intensity images at both sides of the pupil plane.

In addition, the set-up uses a Red Maltese Cross as the fixation target and a chinrest where the patient must be placed. Then, the position of the patient’s pupil is controlled by another camera, allowing the operator to adjust the apparatus until it is centered and focused.

### 2.2 Patients and Measurement Protocol

The sample used in this paper has been described in our previous study [20]. The measurements were performed at the Hospital Universitario Fundación Jiménez Díaz (Madrid, Spain), obtaining a control cohort group with no ocular diseases and a keratoconic group of patients diagnosed and graded by three experienced ophthalmologists. The Hospital Institutional Review Board approved the study protocol that met the tenets of the Declaration of Helsinki. Before t·eyede measurements, informed consent was obtained from the subjects after the nature of the study explanation. The exclusion criteria were presenting other ocular pathologies, being under the legal age, currently pregnant or breastfeeding, or participating in another interventional study within 30 days before starting this study.

Hence, the sample is divided into two main groups: the control group is composed of 43 healthy eyes of 25 patients, and the study group has 43 keratoconic eyes of 27 patients. The demographic characteristics of these are summarized in Table 1.

**Table 1.**
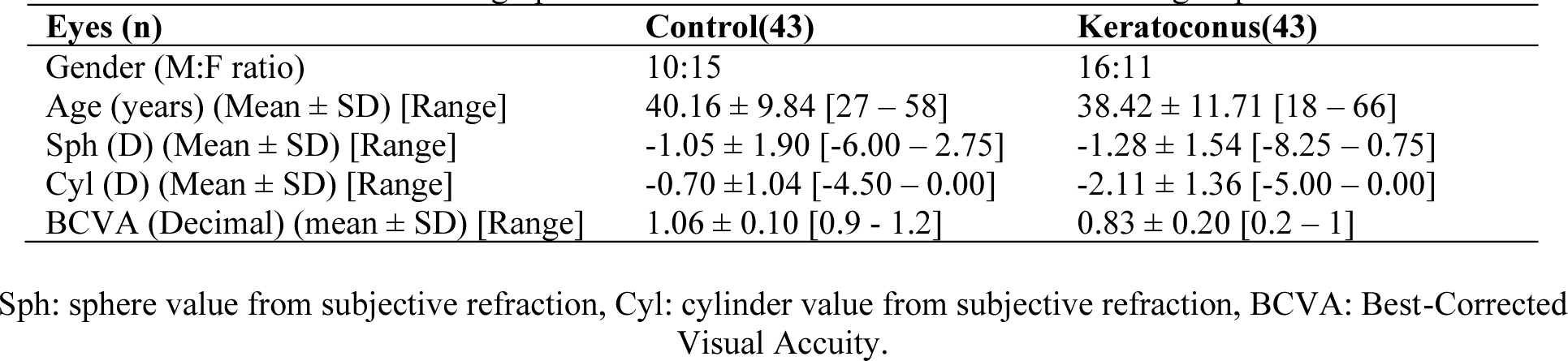
Demographic characteristics of control and keratoconus groups.

| **Eyes (n)** | **Control(43)** | **Keratoconus(43)** |
| --- | --- | --- |
| Gender (M:F ratio) | 10:15 | 16:11 |
| Age (years) (Mean ± SD) [Range] | 40.16 ± 9.84 [27 – 58] | 38.42 ± 11.71 [18 – 66] |
| Sph (D) (Mean ± SD) [Range] | -1.05 ± 1.90 [-6.00 – 2.75] | -1.28 ± 1.54 [-8.25 – 0.75] |
| Cyl (D) (Mean ± SD) [Range] | -0.70 ± 1.04 [-4.50 – 0.00] | -2.11 ± 1.36 [-5.00 – 0.00] |
| BCVA (Decimal) (mean ± SD) [Range] | 1.06 ± 0.10 [0.9 – 1.2] | 0.83 ± 0.20 [0.2 – 1] |

Sph: sphere value from subjective refraction, Cyl: cylinder value from subjective refraction, BCVA: Best-Corrected Visual Acuity.

The division of the keratoconus group between stages has been carried out based on the Pentacam Topographic Keratoconus Classification (TKC), which was corroborated by the criteria of the physicians (the decisive criteria). Only one eye classified as Healthy by the TKC was diagnosed as keratoconus concerning the ophthalmologist criteria (so this eye is included in the KC group), 5 eyes were classified as ‘Suspect’ but the clinician confirmed keratoconus diagnosis, 12 eyes were labeled as stage I, 21 eyes as stage II, 2 eyes as stage III, and last 2 eyes as stage IV.

Regarding the measurement protocol with this aberrometer, the patients had their ophthalmological consultation considered by the clinician. Then, if the subjects matched the keratoconus diagnosis or had no ocular conditions the clinicians explained the study to them, and they were voluntarily invited to sign the informed consent form that allowed the t·eyede operator to take the intensity images. After the image acquisition, the patients are invited to leave the hospital with their ophthalmological consultation concluded.

### 2.3 Image processing

Once the images are acquired, the WFPI software provides the high-resolution raw ocular phase map on the patient’s natural pupil. With this information, a first phase map is displayed up to the 10th Zernike order without the coefficients of piston, tip/tilt, and defocus (named as low-order map), as seen in Fig. 1(a). At the same time, another processed map represents the phase resulting from the subtraction of the raw phase and the low-order phase (named as extremely high-order map), as shown in Fig. 1(b).

Subsequently, the software applies a Gaussian high-pass filter to the extremely high-order map to enhance the high frequencies of the phase (labeled as high-pass filter map), allowing to uncover the hidden details of the low-order map as seen in Fig. 1(c), obtaining for the first time this kind of map in the ocular wavefront analysis thanks to the high resolution of the sensor^19^. This high-pass filter is applied in the frequency domain using a mask for removing frequencies lower than the cut-off value of 25 µm considered as the fittest proved value that sharply enhanced the details contained in this map.

### 2.4 Quantitative Evaluation of High-Pass Filter Mapping

Since the WFPI technique has already been demonstrated to work reliably using the low-order phase to differentiate between healthy and keratoconic eyes^20^, this study will quantitatively characterize the high-pass filter map to show their impact on the decision to diagnose keratoconus as well as its stage. To achieve this purpose, we first take a section of this map (160x160 px for all the eyes) and calculate its Root Mean Square (RMS) and Peak-to-Valley (PV) distance values.

Then, we obtain the FT of this section to analyze its frequency spectrum. The Fourier Transform is a mathematical tool that aims to convert image contents from the ‘spatial’ domain into ‘frequency’ by decomposing this information into a superposition of harmonic functions along the horizontal and vertical axes. For example, if the input image contains a periodic pattern that can be described by a single harmonic, the FT would represent an output image with a specific value of intensity (corresponding to that harmonic) in a gridding scale where its location indicates its frequency value along the x and y axes. Hence, lower frequencies are closer to the origin of the grid (associated with thickness patterns in the spatial domain), while higher frequencies (associated with thinness patterns in the spatial domain) are further away^23–25^. In this analysis, we will extract the amplitude of the predominant frequency in both healthy and keratoconic eyes, as well as for the different disease stages.

The next step is quantitatively assessing the banding patterns using this tool by extracting its spatial period and orientation, as can be seen in Fig. 2. The period (*p*) of the spatial light-dark bands (from the beginning of the light band to the end of the dark band) is obtained knowing the coordinates of the predominant frequency (*fX, fY*) and calculated by the equation (1)^25^; meanwhile the orientation of these bands (*θ*) is calculated as perpendicular to the predominant frequency direction on the Fourier Transform (*φ*) achieved by the expression (2)^25^. In addition, the left eyes have been mirror-symmetry reflected concerning the vertical axis in the spatial domain to compare them against the right eyes.

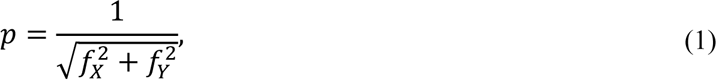

**Fig. 2.**
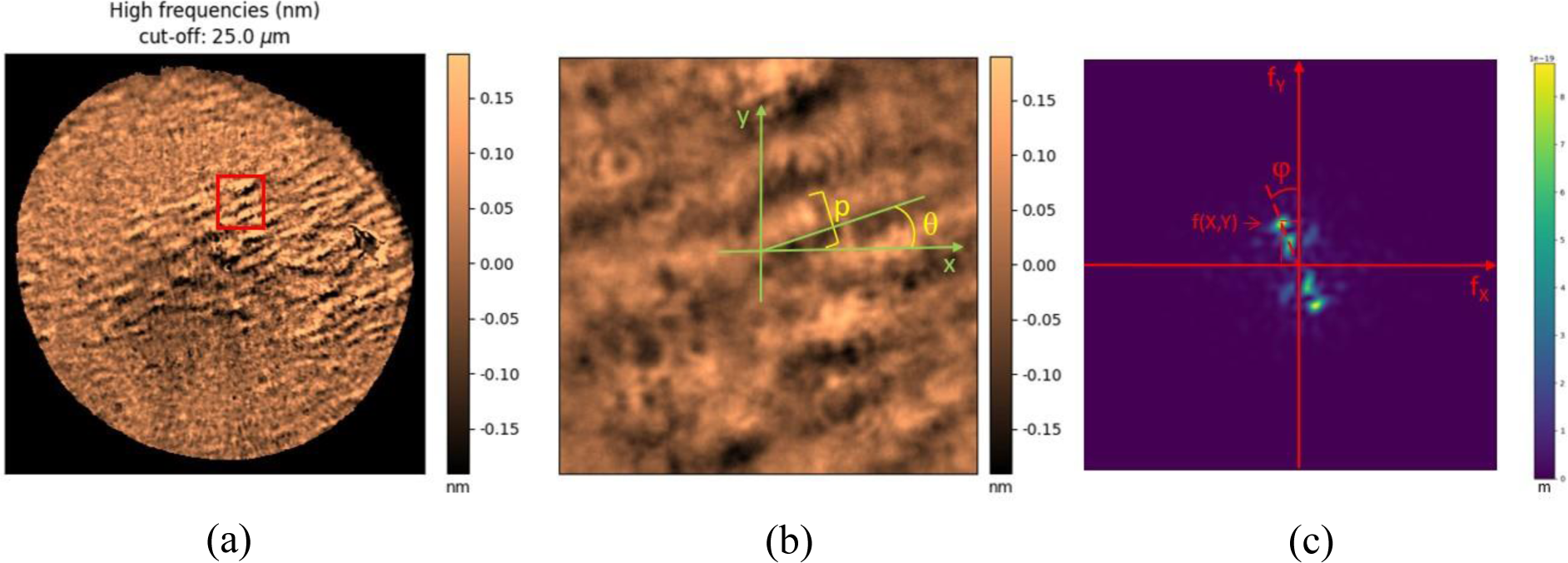
Quantitative assessment of the banding pattern: (a) High-pass filter map of a keratoconic eye with the evaluated section highlighted in red, (b) its magnification, and (c) the Fourier Transform of this section.

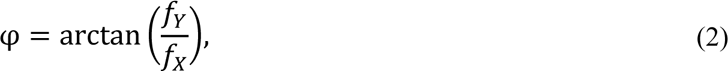

### 2.5 Data analysis

The programs Excel (Microsoft Office 365, v.2301) and Python (v3.9.11) have been used to process and assess the data. The mean and standard deviation values were calculated for all parameters. The statistical analysis has been applied to both main groups using the Shapiro-Wilk test to corroborate the normal distribution of the samples, as well as the Mann-Whitney test to compare both groups. The statistical significance has been established at p values < 0.05. Nevertheless, due to the low number and inequality of the sample in the severity stage groups, this step has not been carried out to compare them.

## 3 Results

### 3.1 High-order phase map quantitative assessment

First, we directly calculated the RMS from the high-pass filter map, whose mean value for the control group was 0.041±0.007 nm (range: 0.025-0.055 nm) and 0.060±0.018 nm (range: 0.036- 0.117 nm) for the keratoconic group. Likewise, the PV mean value obtained for the control group was 0.20±0.03 nm (range: 0.12-0.27 nm), and for the keratoconic group was 0.29±0.09 nm (range: 0.18-0.59 nm). The Shapiro-Wilks test for both parameters in both groups revealed that the control sample had a normal distribution (p > 0.1) while the keratoconic one did not (p < 0.001). Therefore, the nonparametric Mann-Whitney U test was performed to compare both groups, obtaining statistically significant differences (p < 0.001) on both RMS and PV.

Although this analysis cannot be applied to the disease stages, the trend for each parameter concerning the grades is depicted in Fig. 3. As can be seen, RMS tends to increase with the severity stage, as PV does.

**Fig. 3.**
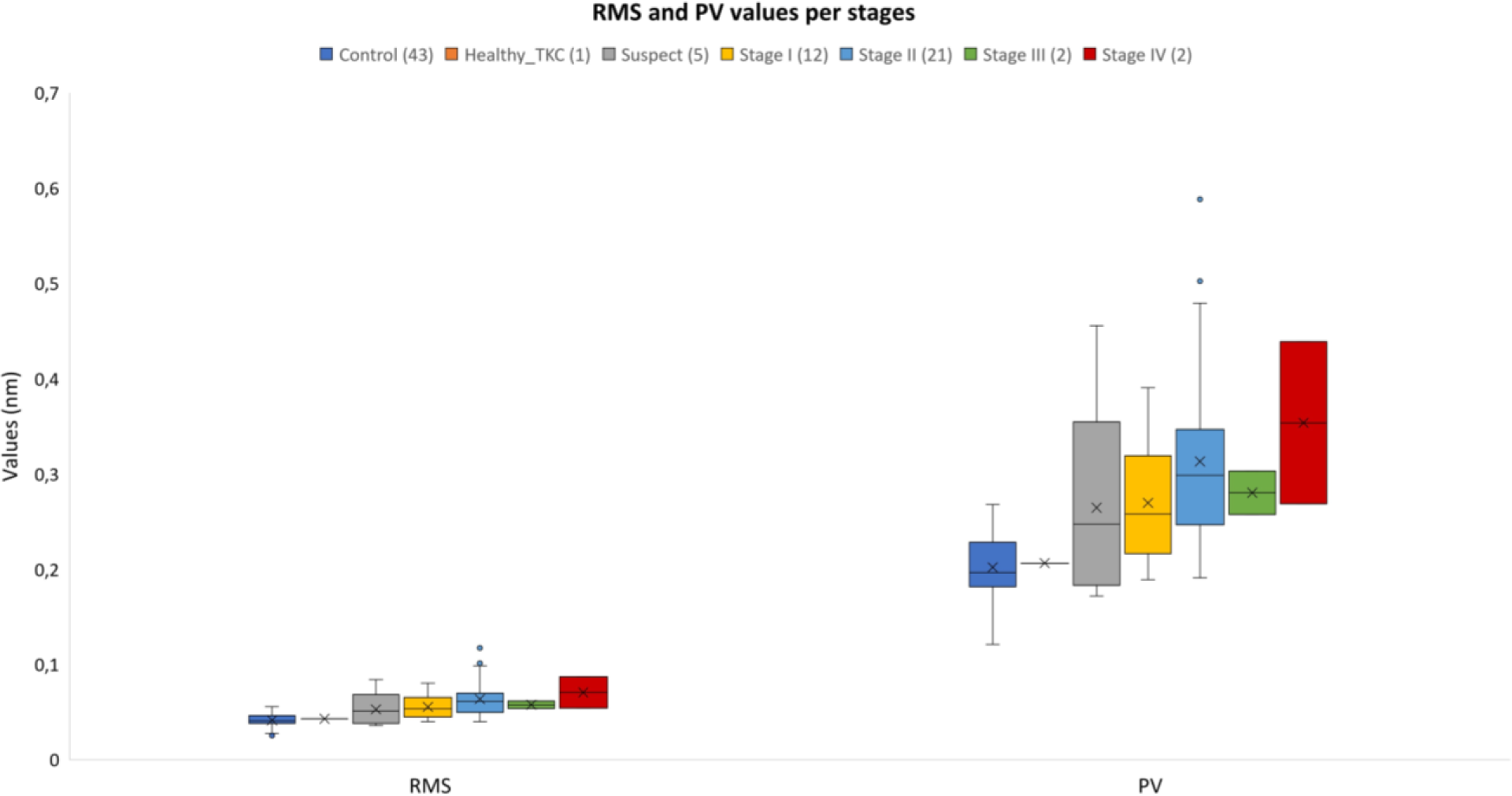
The RMS results for each stage are represented on the left side, while the PV results are depicted on the right side.

Regarding the amplitude of the predominant frequency obtained with the FT, it reached the value of 4.39e-12±3.66e-12 nm (range: 6.12e-13-2.01e-11 nm) for the control group and 5.97e- 11±8.91e-11 nm (range: 1.77e-12-5.22e-10 nm) for the keratoconic group. Performing the same statistical analysis, in this case, the samples did not have the normal distribution according to the Shapiro-Wilks test (p < 0.001). Therefore, the Mann-Whitney test was carried out to compare both groups, obtaining statistically significant differences between them (p < 0.001). At the same time, the comparison has been made between stages to conclude that the amplitude value increases according to the severity of the disease, as shown in Fig. 4.

**Fig. 4.**
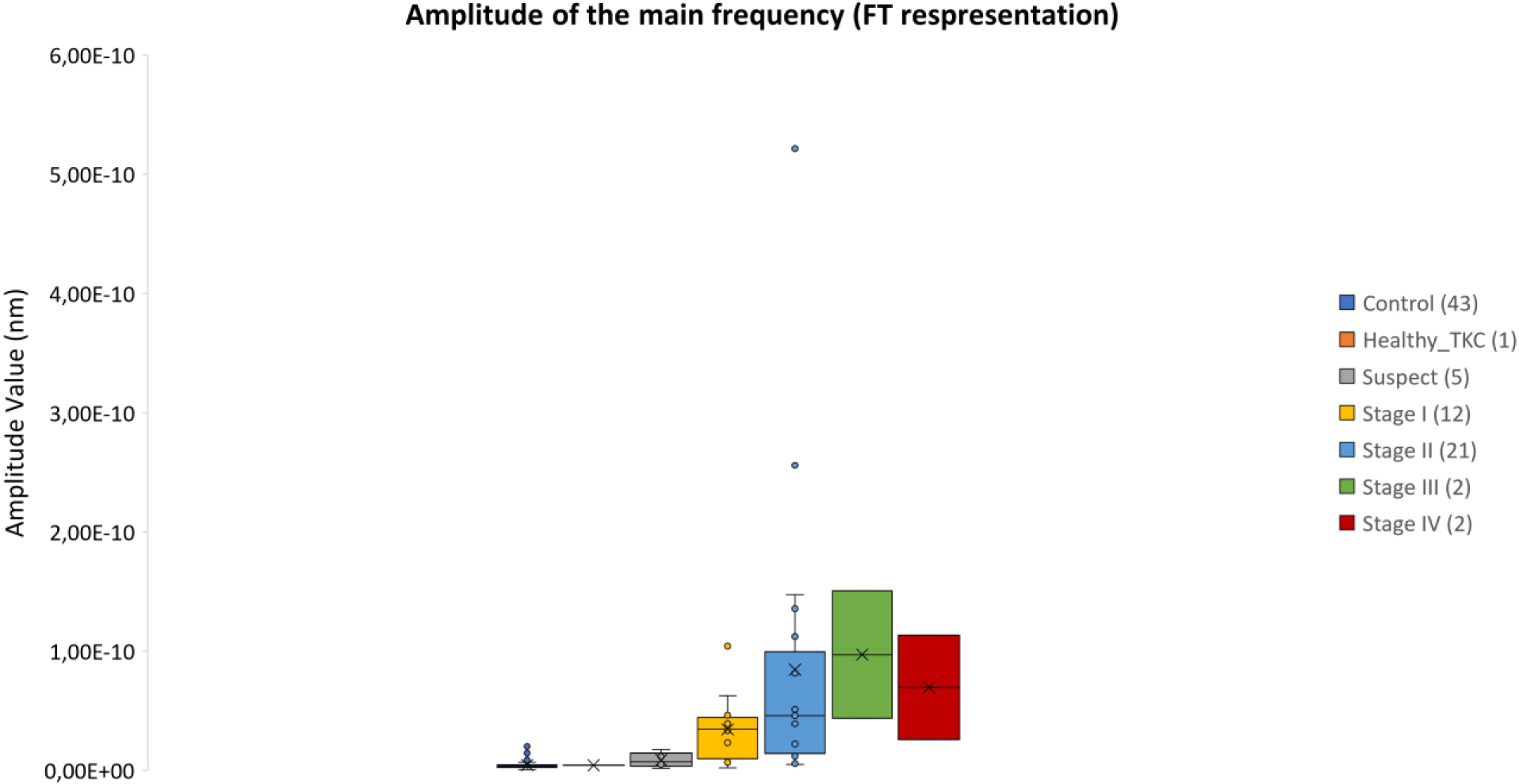
Representation of the amplitude tendency between stages.

### 3.2 Quantification and analysis of the banding pattern

From a total of 43 eyes with keratoconus, 33 of them had banding pattern (2 suspects of keratoconus, 8 with stage I, 19 with stage II, 2 with stage III, and 2 with stage IV), and only 2 from the 43 healthy eyes. Thus, almost 35 eyes presented a banding pattern, divided into 18 right and 17 left eyes.

Regarding the periodicity of the banding pattern, the mean obtained for the total eyes (35) was 51.4±9.4 µm (range: 30.5-71.9 µm); for the right eyes (18) was 51.5±11.5 µm (range: 30.5-71.9 µm); and for left eyes (17) was 51.4±6.6 µm (range: 43.4-63.4 µm). If the sample is subdivided into severity stages, the healthy eyes (2) presented a mean value of 55.5±13.9 µm (range: 45.7- 65.4 µm), the suspect eyes had a mean value of 66.40±7.91 µm (range: 60.8-71.9 µm), the mean for the stage I eyes (8) was 49.4±6.8 µm (range: 38.5-58.4 µm), for the stage II (19) it was 51.4±8.1 µm (range: 38.1-65.1 µm), for the stage III (2) the mean result was 55.7±15.5 µm (range: 44.7- 66.7 µm), and for the stage IV (2) was 37.6±10.1 µm (range: 30.5-44.7 µm).

On the other hand, the orientation results were evaluated in the range of 0 to 180°, as can be seen in Fig. 5. Dividing the total sample between the right and left eyes (Fig. 5(a)), the values for the right eyes (red spots) were concentrated in the oblique angles around 20° and 135°; otherwise, the values for the left eyes (blue squares) were scattered around 20° and throughout the rest of the range. In other words, the preferred orientation in both eyes was more oblique, tending to horizontal rather than vertical.

**Fig. 5.**
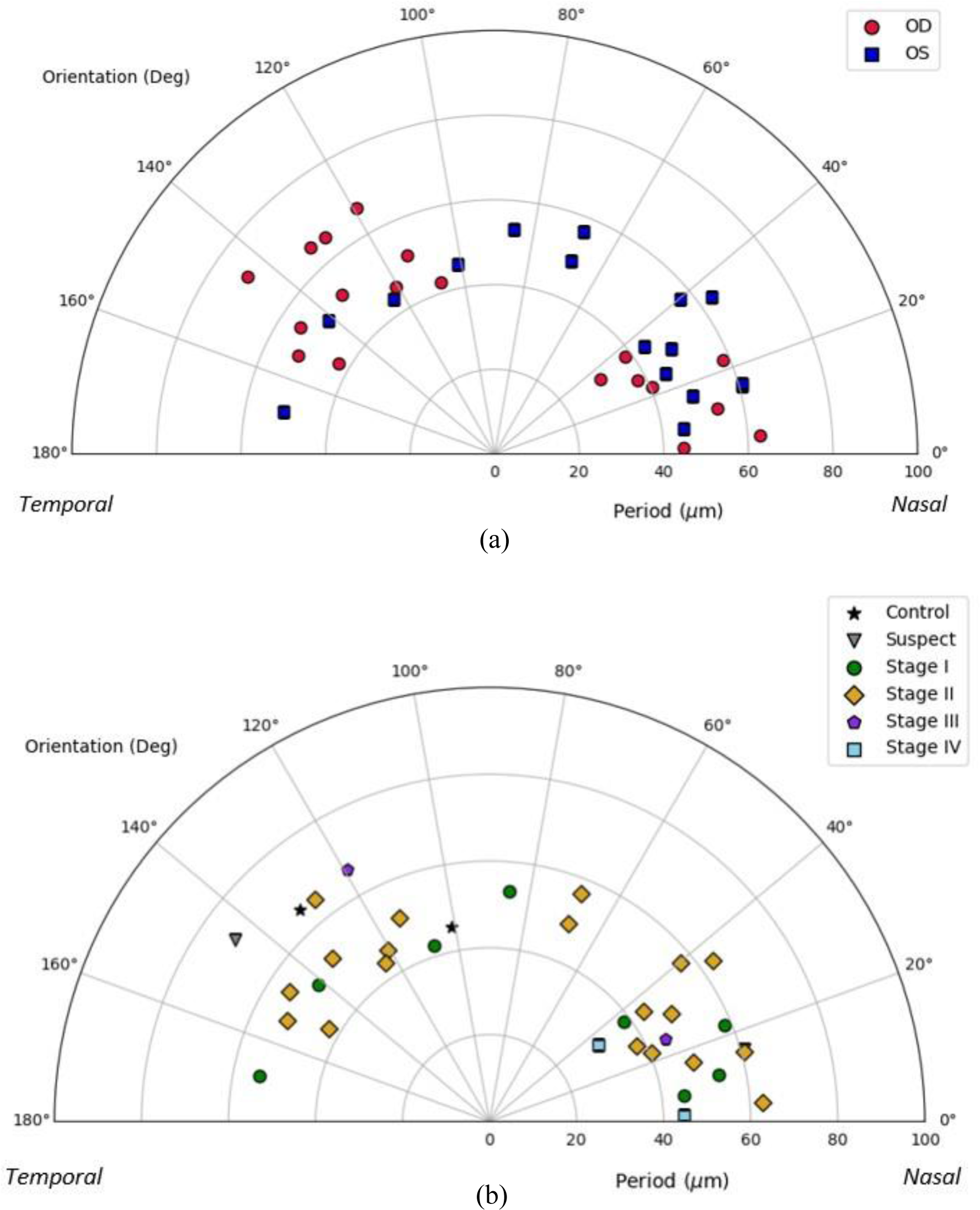
Orientation and period values of the banding pattern for all the eyes: (a) Comparison between eyes (OD) (red spots) and left eyes (OS) (blue squares), and the (b) comparison between stages: Control (black stars), Suspect (grey triangles), Stage I (green spots), Stage II (yellow diamonds), Stage III (purple pentagons), Stage IV (sky blue squares).

Comparing these results by stages (Fig. 5(b)), for the two healthy eyes (black stars) the values were 101° and 132°, while the suspect cases (grey triangles) presented the fringes at 144° and 16°; four eyes in stage I (green spots) were obliquely positioned at approximately 20°, while the other four were dispersed throughout the rest of the angles; in stage II (yellow diamonds), nine eyes were embedded along the first 40° degrees, two orientated more obliquely at 70° approximately, and the rest of values were accounted between 115° and 150°; in stage III (purple pentagons) the two values were 119° and 25°; and at least the two eyes of stage IV (sky blue squares) had the values of 35° and 2°.

From these results, we can summarize that the period of the pattern is independent of the laterality of the eye and the stage because the values are embedded in the range of 40 to 60 µm as the orientation is commonly oblique regardless of the eye or severity.

## 4 Discussion

In this study, we quantitatively evaluated the high-pass filter map obtained with t·eyede in a sample of both healthy and KC eyes. This specific map has been presented for the first time in the work of Bonaque et al. in a small sample with healthy eyes where each eye has different “ocular structures” (the details revealed), but there is no specific pattern^19^. However, in the subsequent study, using the initial prototype of this device to compare the aberrometry between KC and healthy eyes, we qualitatively found a characteristic pattern in most of the maps corresponding to keratoconic eyes such as dark-light alternating bands, concretely in 76.74% of this sample, meanwhile in the rest might not be detectable due to the early stage of the disease or the great decentration of the cone concerning the pupil center^20^. These bands are detectable due to the effect that they cause on the ocular phase, as they have never been seen before using the wavefront measuring technique because no other sensor had sufficient resolution to uncover them.

Although this event has not been observed in phase studies, several papers on keratoconic corneas inform about it using other devices such as slit-lamp microscopy^26^, light microscopy^27^, confocal microscopy^27–30^, and second harmonic generation^31–35^. There is no unique name for this pattern because some authors refer to it as Vogt’s striae^27,28^, folds^30^, or banding pattern^36–38^. But all of them describe it as a phenomenon produced by the stress suffered over the stromal collagen lamellae due to the outgrowth of the cone, and this event appears in almost 50% of their samples, which agrees with our percentage. One of the limitations of this study is the fact that t·eyede measures the total ocular phase and not of a single ocular structure as the cornea, so we cannot conclude with certainty that the pattern obtained in the high-pass filter map corresponds to folds created in the stroma by corneal stress and is an isolated phenomenon of the Keratoconus disease. Then, it would be interesting for future works measuring post-surgery corneal ectasia, Pellucid Marginal Degeneration, or crosslinked-treated corneas (pre- and post-treatment). To solve this disadvantage, a prototype is currently being developed capable of measuring the phase of different ocular structures (tear, cornea, crystalline lens, etc.) and corroborating where these bands came from.

Moreover, we decided to quantitatively assess the bands found in the high-pass filter map with the Fourier Transform because it fits adequately in the study of periodic patterns like this, and many studies applied it to analyze collagen structures from different tissues^21–24^. Nevertheless, there is no unique method to assess this kind of pattern^31,33,35^, and one of our following goals will be to compare Fourier methods with others, such as the Radon Transform, which is also capable of automatically detecting and quantifying the band patterns^39^. We also plan to make use of Artificial Intelligence to label automatically and individually each of the bands for better quantification of the problem and subsequent classification between those subjects who presented this pattern and those who did not, as previous authors had worked with this tool with topography and tomography descriptors to diagnose and evaluate the studied disease^40–44^.

In the presented work, the period of the bands obtained in the FT has a mean of 50 µm with a 95% Confidence Interval of 46 to 54 µm, which is associated with the width of the bands. If we go back to the previous references, these bands have been qualitatively assessed^28–31^, and only Morishige et al. quantify them using confocal microscopy examination of four keratoconic eyes, revealing values along the thickness of the stroma closer to our period values^35^. Many authors discussed that the collagen lamellae of the posterior stroma run straight and parallel in vertical orientation and have the appearance of ‘boarding sheets’ embedded in the range of 100-200 µm, but the lamellae of the anterior stroma have thinner width and their orientation can be found as horizontally as vertically^28,45^. Hence, in the hypothetical case that our detected bands would be collagen lamellae, we cannot assure which section of the stroma is affected, but we are aware that it induces changes in the ocular wavefront phase. Mazzotta et al. concluded that the detection of such patterns, with or without Vogt’s striae, could represent a relative contraindication to perform a riboflavin-UVA-induced corneal cross-linking due to the appearance of stromal haze that could lead to stromal scarring^38^. Although this assumption should be corroborated, and we do not know the severity of the sample and how this pattern is affected, we thought that t·eyede is capable of detecting accurately the early apparition of these bands and could be useful to apply the fittest treatment according to that sign and other critical parameters.

On the other hand, in our study, the mean orientation was oblique rather than vertical, as part of the literature described^27–30,37^. Hollingsworth and Efron attached a diagram (Ref. 27, Fig. 9) of possible orientations of the stress patterns of a keratoconic left eye that are not just vertical (applicable mirror symmetrically for the right eye), and this simulation agrees with the orientations that we saw. Moreover, in the works of Tan et al. and Meek et al. the researchers observed that the alterations of the stromal collagen induce formations of “collagen bundles” centripetally disposed around the cone apex, allowing the assessment of the corneal biomechanical response in the weakest zone (the cone apex)^46,47^. In addition, Mazzotta et al. showed in their observations with confocal microscopy similar patterns with vertical, horizontal, reticular, and oblique orientation also in patients without clinical evidence of Vogt’s striae (as most of our subjects), reporting that these bands appear in the anterior, intermediate, and posterior stroma and would be associated with advanced progressive keratoconus^38^, which coincides with our cases with clear evidence of this sign. Moreover, as we presented in the previous paper, some patterns tend to bend even parallel to each other unless they run straight like Vogt’s striae (Ref. 20 Fig. 3 (e) – (f)), as the images that are shown in the studies with an ex vivo examination using Second Harmonic Generation^31,32^ or confocal microscopy^34^.

Regarding the quantitative analysis of the high-pass filter map, we calculated the RMS and PV of a section to reach other parameters that can help in the KC detection, obtaining for both metrics results statistically different between the control and the keratoconic group as we presented in the previous study with the RMS of astigmatism, coma, and HOA^20^. For further studies, we will consider the sample size, because the sample presented was small and unequal between disease stages, so we cannot compare the marked suspect cases with the controls or compare severe stages (III and IV) with the mid-stages (I and II) where we have more sample. Nevertheless, in our case, we saw that the values of RMS and PV tend to ascend according to the severity of the disease. For that reason, we will consider this for further studies, as we think that these parameters combined with the others obtained by t·eyede measurement and processing (as the amplitude of the predominant frequency in FT representation, astigmatism, and coma), may be sufficiently accurate to distinguish between normal or pathologic eye also between stages even if no banding is presented.

Therefore, banding patterns have been possibly observed with the current phase measurement method when they are not visible at first glance with the slit lamp on the basis that doctors only referenced 5 keratoconic eyes with “streaks” from the 43 evaluated. Along with this, these bands can be quantified objectively using the Fourier Transform as a reliable tool, which automatically performs the analysis without relying on any operator criteria, and the distribution of the energy is appreciated clearly in its representation between healthy and keratoconic eyes, as we try to explain visually in Fig. 6. Here is presented the high-pass filter map of a healthy (Fig. 6(a)) against two keratoconic eyes with different stages (Stage I in Fig. 6(b); and Stage II in Fig. 6(c)), as their respectively Fourier Transform representation (Fig. 6(d)-(f)). Here, it is possible to see that the energy on the healthy eye is scattered along a circumference without any tendency, and the scale bar of the frequencies’ amplitude displays values much lower than the other two scales. Whereas, in the representations of the keratoconic eyes, the energy is concentrated in two symmetrical lobes, which represent the predominance of the bands (with the perpendicular orientation of the lobes) regarding the rest of the details of the map, as well as the increase on the scale bar values in the most severe stage.

**Fig. 6.**
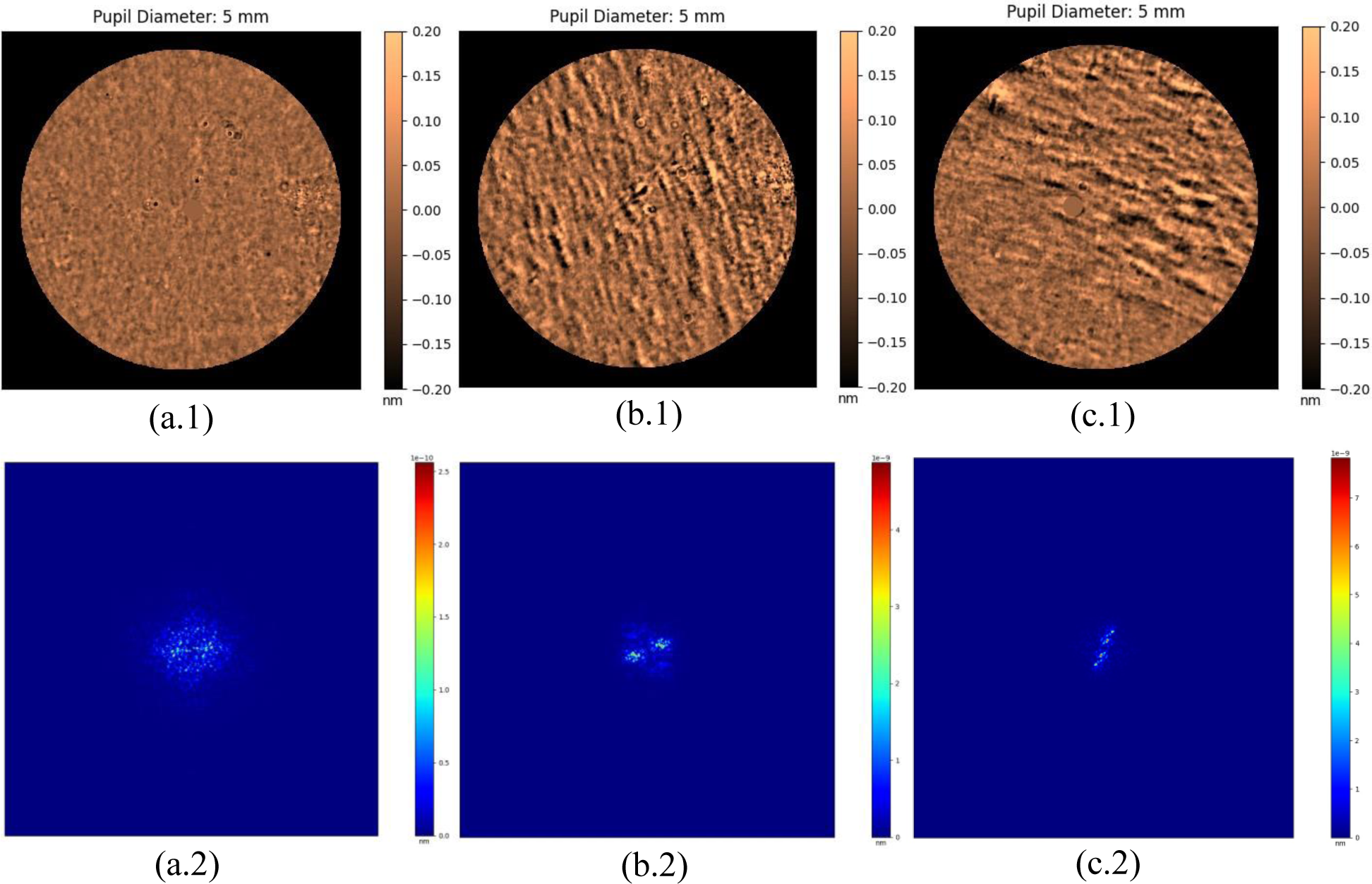
Examples of (a) a right healthy eye, (b) a right Stage I keratoconic eye, and (c) a left Stage II keratoconic eye

In summary, the literature refers to a pattern of dark-light bands that appear in the keratoconic cornea due to the stress suffered by the stromal collagen lamellae by the crescent protrusion of the cone, which are detectable on the moderate and several cases with the slit-lamp, confocal microscopy, and second harmonic generation examination. We observed a similar pattern using a high-resolution ocular aberrometer after processing the extremely high orders of the whole phase, reaching statistical differences between healthy and pathological eyes. Then, we can characterize this pattern using the Fourier Transform, obtaining results closer to those described in the bibliography regarding width and orientation.

## 5 Conclusions

As closure, we demonstrate that the high resolution achieved by the t·eyede aberrometer can quantitatively discern between healthy and keratoconic eyes and be the one in revealing a specific banding pattern on keratoconic eyes through the phase information. These bands can be quantitatively determined using the Fourier Transform, which can accurately characterize them by their orientation and period, obtaining that most of these bands are oblique rather than vertical and have a width of approximately 50 µm. Further clinical studies with this device are needed to confirm that the presence of these bands is due to the stress suffered by the stroma during the cone formation, to understand the corneal biomechanics, and to corroborate how this event may contribute to refining the diagnosis process, monitoring, and fitting the treatment of the keratoconus.

## Code, Data, and Materials

The data that support the findings of this article are not publicly available, but can be requested from the author at.

## Data Availability

All data produced in the present study are available upon reasonable request to the authors

## Acknowledgments

We would like to thank Dr. José Gil Marichal Hernández for his advice and evaluation of the quantitative analysis.

